# ‘I need policymakers to listen’: a stakeholder needs assessment concerning anticipatory injectable medication systems, using a novel qualitative survey

**DOI:** 10.64898/2026.01.02.26343333

**Authors:** Binu Perera, Ben Bowers

**Affiliations:** University of Cambridge Department of Public Health and Primary Care Forvie Site, Robinson Way, Cambridge, UK CB2 0SR; University of Cambridge, Engineering Design Centre, Department of Engineering, Trumpington Street, Cambridge, UK CB2 1PZ

**Keywords:** Anticipatory prescribing, anticipatory medications, palliative medicine kit, injection, terminal care, palliative care, qualitative methods, end of life care, home palliative care

## Abstract

**Background:** Anticipatory injectable medications for symptom control are a key end-of-life care intervention. However, ensuring their safe and timely use in the community is a global challenge. The needs and priorities of stakeholders involved in processes for prescribing and administering these medications remain underexplored. We must understand these perspectives to design inclusive and adaptive systems.

**Aim:** To identify the needs and priorities of key stakeholders involved in community-based systems for using anticipatory injectable medications.

**Design:** We adopted a qualitative exploratory design, using an online survey between September and October 2024. Participants provided anonymised demographic information and completed up to four prompts capturing their stakeholder role, needs and priorities. Data were analysed using a combined inductive–deductive framework to produce synthesised shortlists of priorities and needs.

**Setting/participants:** UK-based professional and public participants were recruited through social media, professional networks, charities, and public engagement events.

**Results:** In total, 439 participants contributed 729 responses across various stakeholder groups. Findings revealed substantial diversity in stakeholder needs and priorities, both within and between groups. However, most stakeholder groups prioritised timely care, minimising of suffering, and wanted nationally consistent guidance for using injectable medications. Broader societal influences also shaped responses.

**Conclusions:** Our findings highlight wide diversity in priorities and needs between stakeholders for using anticipatory injectable medications in the community. We propose that inclusive system design should include comprehensive assessment of key stakeholders’ needs and priorities, with the aim of providing better care. Our study demonstrates that stakeholder needs assessment offers a valuable framework to achieve this.

**What is already known about the topic?:**

- Anticipatory injectable medications are a widely used intervention in several countries to support timely end-of-life symptom control at home.
- There are ongoing challenges with delays, inconsistent access, and variations in prescribing and governance across regions, indicating that system design influences both timeliness and safety.
- Existing research has primarily focused on the needs of individual professional groups, and no prior work has mapped the differing needs of all stakeholders involved in these systems.

**What this paper adds?:**

- Our study demonstrates that stakeholder groups have diverse needs but most share some core priorities -timely care, national consistency in practice guidance, and minimising suffering.
- Wider societal factors and concerns shape stakeholder expectations of end-of-life medication systems.
- Our approach to stakeholder needs assessment reveals system requirements that consensus-based or single-perspective approaches often overlook.

**Implications for practice, theory, or policy:**

- System improvements should be tailored to the specific needs of key stakeholder groups rather than assuming uniform priorities.
- Strong cross-stakeholder support exists for national, practical guidance on anticipatory prescribing, equipment, training, and governance.
- Stakeholder needs assessment offers a useful method for designing safer, more responsive end-of-life medication systems.

## Introduction

Over half of all deaths globally are estimated to take place at home.^1^ Community healthcare systems need to respond effectively to the variable symptom management needs of patients in their last weeks and days of life. However, dying at home comes with several challenges around providing safe, effective and timely symptom control. Anticipatory injectable medications are an important part of this care internationally, including in the United Kingdom (UK), Canada and Australia.^2^ These medications are prescribed in advance of anticipated need for common symptoms including pain, respiratory secretions, nausea and vomiting, and agitation^3^. They are stored in the patient’s home with a permission to administer chart (‘standing order’), to be administered by visiting nurses and paramedics when needed. ^4–6^

This study is part of a wider research programme investigating the factors involved in the safe, effective, and timely use of anticipatory end-of-life medications at home. We investigated the priorities and needs of key actors involved in community-based systems for using anticipatory injectable medications via a ‘Stakeholder Needs Assessment’ – a novel technique used in engineering design.^4^ This approach is rooted in an inclusive design ethos, where meeting the diverse requirements of key stakeholders produces better designed systems for all.^4,5^ We aimed to identify areas of consensus and differing perspectives within and across stakeholder groups, to define their top priorities and needs and guide future system (re)designs.

## Aims

To identify the needs and priorities of key stakeholders involved in community-based systems for using anticipatory injectable medications.

## Methods

### Study design

We conducted an online qualitative survey of participants living and working in the UK, using Qualtrics. The study was approved by the Department of Engineering Research Ethics Committee, University of Cambridge [reference: 539, 2024].

### Recruitment and data collection

We recruited participants via social media (including X and Bluesky), public engagement events and professional conferences between 23 September and 4 November 2024. All participants provided informed consent and anonymised demographic data. Participants completed the following short statements:^4^

> *As a… [stakeholder type]*
>
> *I need… [free text]*
>
> *So that… [free text]*

Participants were able to complete the statement up to four times, providing data about individuals’ needs and priorities in the various roles they may occupy. This novel approach enabled us to canvas a wide range of needs and priorities concisely, across a variety of stakeholder groups.

### Data analysis

We conducted an inductive and deductive framework analysis^6^ to identify common and divergent perspectives. The responses were transferred into NVivo (Version 14) and coded by B.P., focused on identifying the priorities and needs within narratives; coding was refined by B.P. and B.B. through reflexive discussions.

Initial codes were aggregated into stakeholder-specific lists, which were then refined into longlists and then shortlists by B.P. and B.B. We discussed two longlists with a group of eight public contributors with lived experience of end-of-life care as family carers or professionals. These discussions informed our iterative process of producing the shortlists: thematic representations of the responses received. The output of our analysis is two stakeholder-specific shortlists per stakeholder group: an ‘I need’ shortlist, which corresponds to system-related desires of each stakeholder, and a ‘so that’ shortlist, which lays out the priorities of each stakeholder group. The ‘I need’ shortlist acts as a list of potential avenues to achieve stakeholder-specific desires ‘so that’. ^4^

## Results

In total, 439 individuals responded to our survey, producing 729 statements. Participant characteristics and a break-down of self-identified stakeholder groups are detailed in Table 1.

**Table 1.**
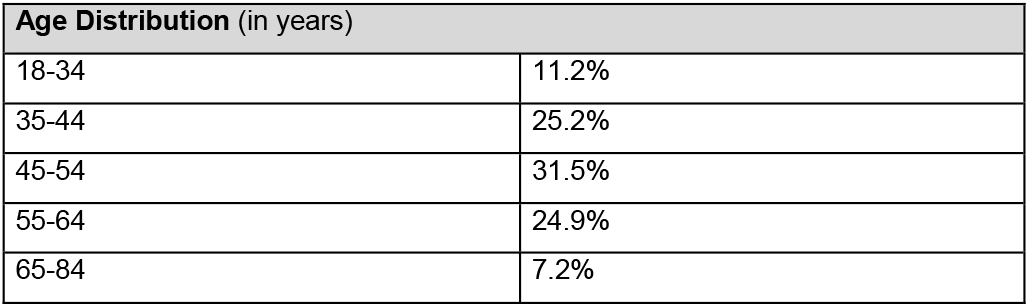
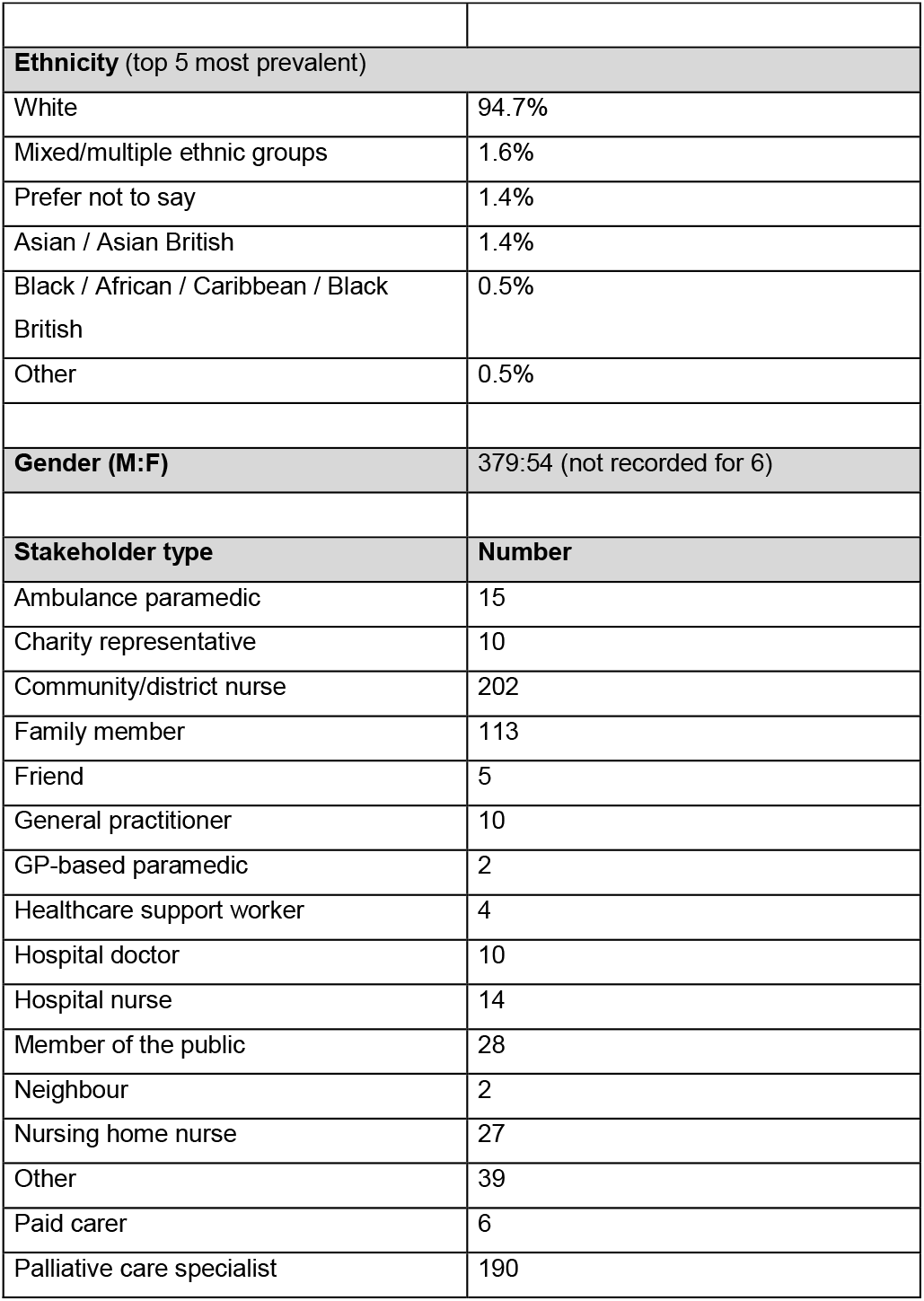
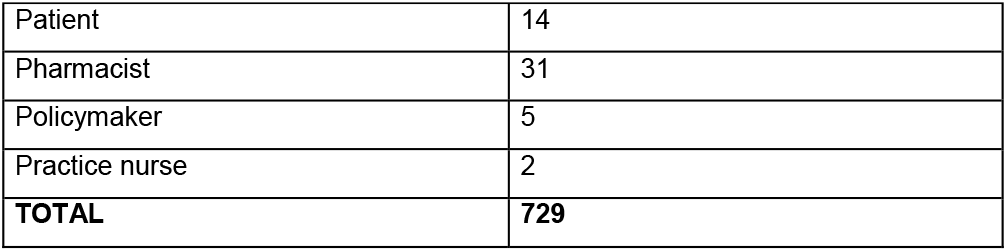
Participants characteristics and stakeholder groups.

| Age Distribution (in years) |  |
| --- | --- |
| 18-34 | 11.2% |
| 35-44 | 25.2% |
| 45-54 | 31.5% |
| 55-64 | 24.9% |
| 65-84 | 7.2% |
| **Ethnicity (top 5 most prevalent)** |  |
| White | 94.7% |
| Mixed/multiple ethnic groups | 1.6% |
| Prefer not to say | 1.4% |
| Asian / Asian British | 1.4% |
| Black / African / Caribbean / Black British | 0.5% |
| Other | 0.5% |
| **Gender (M:F)** | 379:54 (not recorded for 6) |
| **Stakeholder type** | **Number** |
| Ambulance paramedic | 15 |
| Charity representative | 10 |
| Community/district nurse | 202 |
| Family member | 113 |
| Friend | 5 |
| General practitioner | 10 |
| GP-based paramedic | 2 |
| Healthcare support worker | 4 |
| Hospital doctor | 10 |
| Hospital nurse | 14 |
| Member of the public | 28 |
| Neighbour | 2 |
| Nursing home nurse | 27 |
| Other | 39 |
| Paid carer | 6 |
| Palliative care specialist | 190 |
| Patient | 14 |
| Pharmacist | 31 |
| Policymaker | 5 |
| Practice nurse | 2 |
| **TOTAL** | **729** |

Figure 1 shows the final ‘priorities and needs’ shortlist for the ‘Nurses’ stakeholder group. The shortlist diagrams for each major stakeholder group can be found in the supplementary material.

**Figure 1.**
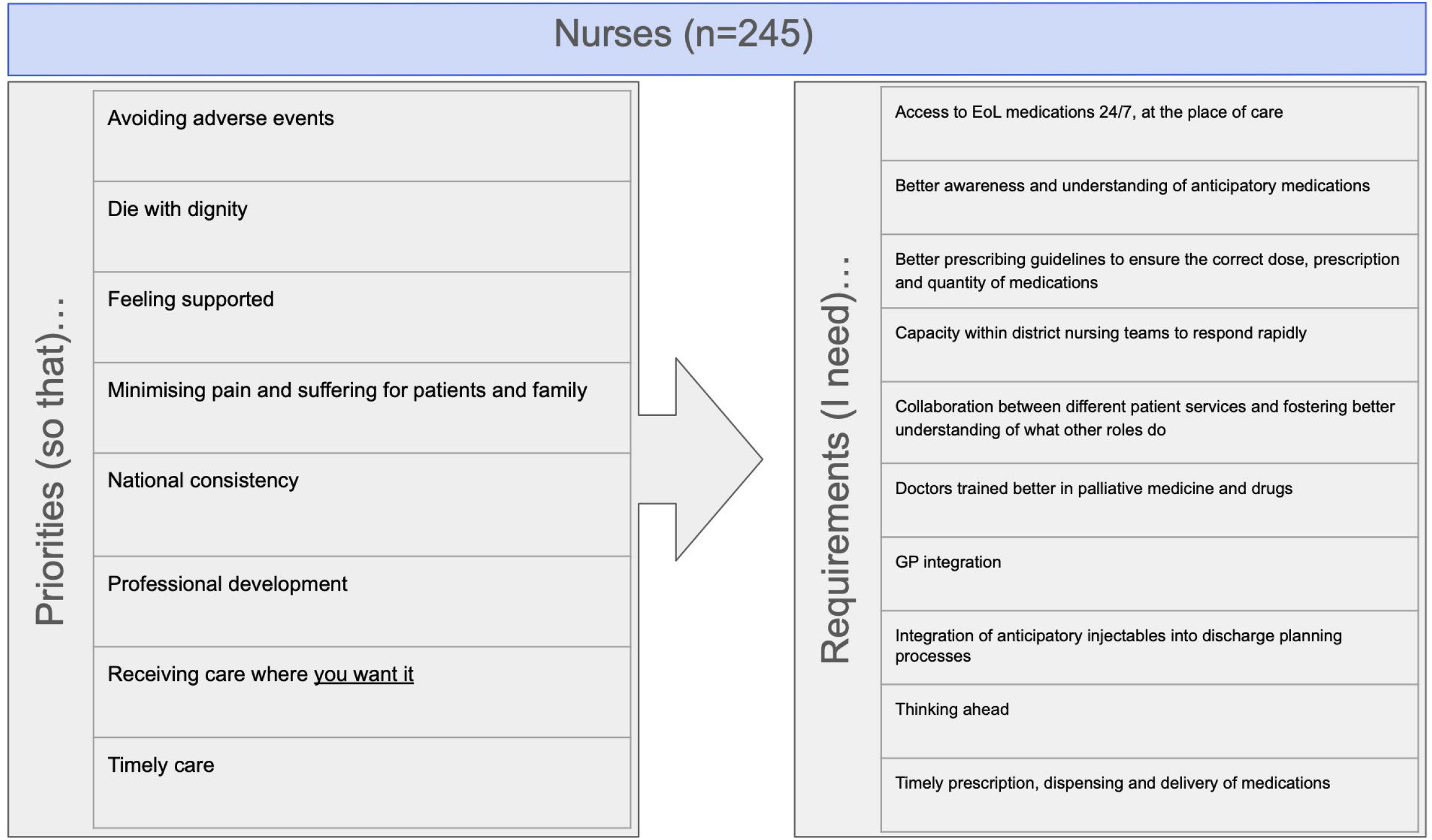
Nurses stakeholder priorities and needs shortlist. ***Figure 1 provided separately as per submission guidelines***. On comparing the different shortlists, there was notable diversity in priorities and needs between different groups. However, themes of ‘access to timely care’, ‘national consistency’, and ‘minimising pain and suffering’ were consistently important between stakeholder groups. For example, one nurse highlighted the importance of access to timely care by emphasising the negative impacts of not having medications available on patients, families and nurses themselves:

> ***As a:***
>
> *Community/district nurse*
>
> ***I need:***
>
> *‘*…*GPs to be more proactive in planning and listening to community nurses’*
>
> ***So that:***
>
> *‘*…*[we can] manage symptoms as soon as [the] patient requires it. Delay in care and inability to access injectables to relieve symptoms puts nurses in a difficult situation, often having to spend time sorting out prescription of injectables and [permission to administer] charts whilst [the] patient is agitated/[in] pain, which in turn massively impacts the family/friend’*

Another respondent summarised the possible benefits to patients, staff, public, and policymakers if there was a nationally consistent approach to using anticipatory injectable medications:

> ***As a:***
>
> *Policymaker*
>
> ***I need:***
>
> *‘*…*there to be crystal clear guidance around [using] syringe drivers…’*
>
> ***So that:***
>
> *‘*…*there is national consistency, to enable staff moving across county borders to transfer knowledge and skills, it helps the public narrative and understanding around end of life medications, staff to feel more confident and competent, reduces time spent for local leaders pulling together extensive guidance […], and improves the overall practice and clinical effectiveness/patient safety of anticipatory medications’*

There were also references to overarching societal concerns throughout the responses, including assisted dying and national resource constraints. For example, one respondent expressed timely home-based injectable medication care was important as an alternative to assisted dying. At the time of our study, the assisted dying bill was being debated in the UK Parliament:

> ***As a:***
>
> *Member of the public*
>
> ***I need:***
>
> *‘systems to enable […] patients and their relatives with regards to palliation at home/in a care home’*
>
> ***So that:***
>
> *‘*…*[those who wish to] die at home can [do so] without their care being compromised by waits or post code lottery and ensure absence of good palliation doesn’t lead the public to think that assisted dying is the only option…’*

Another respondent referenced the resource constraints UK National Health Service (NHS) services were under, and suggested alternative approaches were needed to ensure timely injectable medication symptom-control care:

> ***As a:***
>
> *Palliative care specialist*
>
> ***I need:***
>
> *‘Patients or carers to be trained in [subcutaneous injectables] administration’*
>
> ***So that:***
>
> *‘*…*[so that] patients don’t have to wait for stretched healthcare services before receiving medications for symptom control’*
>
> These themes reveal how broader social forces and anxieties about pressures on current care systems inform stakeholder priorities. They emphasise the need to consider these wider dynamics in the design and delivery of palliative care.

## Discussion

Our stakeholder needs analysis reveals the significant diversity in priorities and needs between and within stakeholder groups. This is an expected outcome of our study and is a particularly important consideration in the context of (re)designing healthcare systems. Multiple actors and healthcare providers are involved in the effective and timely use of injectable medications^2,6,7^. It is important to recognise the differing priorities and needs when designing these safety-critical systems.^2,7–9^ Even when priorities converge, differing needs mean that cross-stakeholder approaches may not deliver all desired outcomes and can perpetuate misunderstandings about the role of injectable medications in care.^7,10^ Our results are also a reminder for international policy makers and service providers to respond to wider societal concerns about local resource constraints and deficits in existing systems of care.^7,11–13^

We have chosen not to produce a single aggregated list of priorities and needs across stakeholder groups. In presenting the different stakeholders’ shortlists, we hope to highlight their differing priorities and needs, and underscore that though these may vary, there should not be inherent importance attached to specific stakeholders requirements. ^4,5^ For example, responses from pharmacists tended to be more process oriented, with particular importance given to managing risk and tackling governance issues. This contrasts with responses from nurses, which were more generally concerned with protecting patients from unnecessary delays and pain. **All stakeholder priorities and needs are important to injectable medication system (re)design**.

National consistency was important to almost all the key stakeholder groups in our study. Considering this, better injectable medication system design should include provision of national, cross-organisational guidance on anticipatory medications, the timing of prescriptions, equipment supplies, their subsequent use and monitoring.

### Strengths and limitations

Responses were aggregated and condensed into easily digestible shortlists by the authors to produce a practically useful outcome of our survey through grouped stakeholder priorities and needs. However, we recognise that a shortlist can never capture the full range of responses, and in the words of a member of the public contributor group, ‘someone has to make a decision’. We found that the public contributors group discussion was an ineffective means of condensing the longlist into a shortlist. Instead, public contributors provided their own additional priorities and needs which we were unable to include in the analysis.

The shortlists produced are reflections of the strength of the stakeholder needs assessment methods, revealing the broad range of key stakeholders and their requirements. This engineering design method is novel, exploratory and experimental, and provides insights into the breadth of requirements rather than depth of perspectives. ^4,5^ In the future, this research method could be enhanced by following up initial stakeholder needs assessment data collection with more in-depth research interviews with a maximum diversity sample of participants.

## Conclusion

Our findings demonstrate the wide diversity in priorities and needs between stakeholders with regards to systems for using anticipatory injectable medications in the community. Within this stakeholder-specific diversity, there are convergent themes which unite the priorities of stakeholders, including national consistency and timely care. Our novel application of stakeholder needs analysis techniques has been successful in canvassing a wide range of opinions, and this approach can readily be applied to understanding other complex systems. Our approach is particularly effective at challenging the potentially counter-productive dogma of looking for census and the common assumption of uniformity between healthcare providers: a group that is often designed to include doctors, nurses and pharmacists.

Future work needs to refine methods for analysing large volumes of qualitative responses through larger studies. Whilst future developments may utilise pattern recognition/artificial intelligence technologies to achieve this, care must be taken to ensure that responses are analysed in a way that is true to their intended meaning.

## Supporting information

Supplementary images

## Data Availability

The anonymised data used in this study may be requested by researchers through contacting B.B., second author and data custodian, and on completion of a data use and sharing agreement.

## Acknowledgements

The authors would like to thank the eight public contributors for their valuable input, insights and advice. James Brimicombe, Senior Data Manager for the Primary Care Unit, Department of Public Health and Primary Care, University of Cambridge, for his help in planning and running this study. James Cantwell, Communications, Engagement and Patient and Public Involvement and Engagement Manager, Primary Care Unit, for all his help in sharing the survey widely.

## Author contributions

B.B. and B.P. designed the study. B.P. carried out the analysis with input from B.B. Both authors contributed to the interpretation of the results and the writing of the manuscript. B.B. is the guarantor. The corresponding author attests that all listed authors meet authorship criteria and that no others meeting the criteria have been omitted. The first author (B.P.) affirms that the manuscript is an honest, accurate and transparent account of the study being reported.

## References

1. Adair T. Who dies where? Estimating the percentage of deaths that occur at home. BMJ Glob Health 2021; 6: e006766.

2. Bowers B, Antunes BCP, Etkind S, et al. Anticipatory prescribing in community endof-life care: systematic review and narrative synthesis of the evidence since 2017. BMJ Support Palliat Care 2024; 13: e612–e623.

3. McGlinchey T, Early R, Mason S, et al. Updating international consensus on best practice in care of the dying: A Delphi study. Palliat Med 2023; 37: 329–342.

4. Clarkson J. Improving Improvement, https://www.iitoolkit.com/Improving_Improvement_1-28.pdf (2022).

5. Elliott C, Deasley P. Creating systems that work: principles of engineering systems for the 21st century. London: Royal Academy of Engineering, 2007.

6. Smith J, Firth J. Qualitative data analysis: the framework approach. Nurse Res 2011; 18: 52–62.

7. Bowers B, Pollock K, Barclay S. Simultaneously reassuring and unsettling: a longitudinal qualitative study of community anticipatory medication prescribing for older patients. Age Ageing 2022; 51: afac293.

8. Bowers B, Gwyn S, Yardley S, et al. Learning from end-of-life injectable medication patient safety incidents in the community: a mixed-methods analysis. Br J Gen Pract 2025; BJGP.2025.0106.

9. Williams H, Donaldson SL, Noble S, et al. Quality improvement priorities for safer out-of-hours palliative care: Lessons from a mixed-methods analysis of a national incident-reporting database. Palliat Med 2019; 33: 346–356.

10. Brown A, Yardley S, Bowers B, et al. Multiple points of system failure underpin continuous subcutaneous infusion safety incidents in palliative care: A mixed methods analysis. Palliat Med 2025; 39: 7–21.

11. Pollock K, Caswell G, Turner N, et al. The ideal and the real: Patient and bereaved family caregiver perspectives on the significance of place of death. Death Stud 2024; 48: 312–325.

12. Collier A, Dadich A, Jeffs C, et al. ‘The palliative care ambulance’: A qualitative study of patient and caregiver perspectives of an ambulance service. Palliat Med 2023; 37: 875–883.

13. Bowers B, Pollock K, Etkind S, et al. ‘We’ve Taken on a More Advanced Clinical Role’: A Multimethod Study of Community Nurses’ Extended Roles in Palliative Care. J Adv Nurs. Epub ahead of print 17 June 2025. DOI: 10.1111/jan.70019.

