## Supplementary images for "‘I need policymakers to listen’: a stakeholder needs assessment concerning anticipatory injectable medication systems, using a novel qualitative survey"

### Family and Public (n=148)

**Priorities (so that)...**

- Avoiding adverse events
- Die with dignity
- Feeling supported
- Get things right first time
- Inclusion in care planning
- Minimising pain and suffering for patients and family
- National consistency
- Receiving care where you want it
- The right medications in place
- Timely care

**Requirements (I need)...**

- Access to EoL medications 24/7, at the place of care
- Better awareness and understanding of anticipatory medications
- Better prescribing guidelines to ensure the correct dose, prescription and quantity of medications
- Clear plans and instructions in place
- Collaboration between different patient services and fostering better understanding of what other roles do
- Doctors trained better in palliative medicine and drugs
- Early recognition of normal end of life changes
- Education and training opportunities for healthcare professionals
- Effective communication, empathy and understanding which includes listening to patients, family and carers
- GP integration
- Thinking ahead
- Timely prescription, dispensing and delivery of medications

### Nurses (n=245)

**Priorities (so that)...**

- Avoiding adverse events
- Die with dignity
- Feeling supported
- Minimising pain and suffering for patients and family
- National consistency
- Professional development
- Receiving care where you want it
- Timely care

**Requirements (I need)...**

- Access to EoL medications 24/7, at the place of care
- Better awareness and understanding of anticipatory medications
- Better prescribing guidelines to ensure the correct dose, prescription and quantity of medications
- Capacity within district nursing teams to respond rapidly
- Collaboration between different patient services and fostering better understanding of what other roles do
- Doctors trained better in palliative medicine and drugs
- GP integration
- Integration of anticipatory injectables into discharge planning processes
- Thinking ahead
- Timely prescription, dispensing and delivery of medications

### Patients (n=14)

#### Priorities (so that)…

- Avoiding adverse events
- Minimising pain and suffering for patients and family
- Realistic expectations
- Timely care

#### Requirements (I need)…

- Access to EoL medications 24/7, at the place of care
- Availability of staff
- Clear guidelines and instructions
- Confidence in other healthcare professionals
- Listening to patients
- Rapid delivery of drugs
- Thinking ahead

### Pharmacists (n=31)

#### Priorities (so that)…

- Avoiding adverse events
- Die with dignity
- Guidance to back up decision making
- National consistency
- Receiving care where you want it
- The right medications in place at the time of need
- Timely care

#### Requirements (I need)…

- Access to EoL medications 24/7, at the place of care
- Availability of research and evidence
- Clear guidelines and instructions
- Education and training
- Ethicolegal support
- Prescribing safeguards
- Syst4ems for medication disposal
- Timely prescription, dispensing and delivery of medications

### Doctors and Specialists (n=210)

**Priorities (so that)…**

- Avoiding adverse events
- Die with dignity
- Feeling supported
- Get things right first time
- National consistency
- Plans are tailored to each patient
- Receiving care where you want it
- The right medications in place at the time of need
- Timely care

**Requirements (I need)…**

- Access to EoL medications 24/7, at the place of care
- Better awareness and understanding of anticipatory medications
- Better prescribing guidelines to ensure the correct dose, prescription and quantity of medications
- Capacity within district nursing teams to respond rapidly
- Collaboration between different patient services and fostering better understanding of what other roles do
- Doctors trained better in palliative medicine and drugs
- GP integration
- Integration of anticipatory injectables into discharge planning processes
- Thinking ahead
- Timely prescription, dispensing and delivery of medications
